# Effectiveness of Rosuvastatin plus Colchicine, Emtricitabine/Tenofovir and a combination of them in Hospitalized Patients with SARS Covid-19

**DOI:** 10.1101/2021.07.06.21260085

**Authors:** Hernando Guillermo Gaitán-Duarte, Carlos Álvarez-Moreno, Carlos Javier Rincón-Rodríguez, Nancy Yomayusa-González, Jorge Alberto Cortés, Juan Carlos Villar, Juan Sebastián Bravo-Ojeda, Ángel García-Peña, Wilson Adarme-Jaimes, Viviana Alejandra Rodríguez-Romero, Steffany Lorena Villate-Soto, Giancarlo Buitrago, Julio Chacón-Sarmiento, Martín Macías-Quintero, Claudia Patricia Vaca, Carlos Gómez-Restrepo, Nelcy Rodríguez-Malagón

## Abstract

**BACKGROUND:** The effectiveness of rosuvastatin plus colchicine, emtricitabine/tenofovir, and of their combined use in hospitalized patients with coronavirus disease 2019 (Covid-19) pneumonia is unclear.

**METHODS:** In each hospital, hospitalized adults with Covid-19 pneumonia, were randomly assigned, in a 1:1 ratio, to receive: a) standard of care; or b) emtricitabine/tenofovir; or c) colchicine + rosuvastatin; or d) emtricitabine/tenofovir + colchicine + rosuvastatin. The primary outcome was all-cause mortality within the first 28 days after randomization. Severe adverse events (SAE) were those with a high probability of being treatment-related.

**RESULTS:** 633 patients were randomized in 6 hospitals in Bogota, Colombia. Overall, 98% of the patients received glucocorticoids during hospitalization. The cumulative incidence of death through day 28 was 10.7% in the emtricitabine/tenofovir + colchicine + rosuvastatin arm, 14.4% in the colchicine + rosuvastatin arm, 13.8% in the emtricitabine/tenofovir arm, and 17.4% in the standard of care arm, with adjusted risk differences (aRD) against the standard treatment of -0.07 (95% confidence interval [CI], -0.17 to 0.04), aRD -0.03 (95%CI: -0.11 to 0.05) and aRD: -0.05 (95%CI: -0.15 to 0.05), respectively. Need for invasive mechanical ventilation was lower in the emtricitabine/tenofovir + colchicine + rosuvastatin arm compared to the standard treatment arm, aRD: -0.06 (95%CI: -0.11 to -0,01), but no differences were found between the other comparisons. SAE occurred in 3 patients distributed in the 3 treatment arms.

**CONCLUSIONS:** Among patients hospitalized with moderate and severe SARS Covid-19, the use of the emtricitabine/tenofovir + colchicine + rosuvastatin combination emerges as a treatment alternative.

ClinicalTrials.gov number: NCT04359095

## INTRODUCTION

Since the description of coronavirus disease 2019 (Covid-19), several effective vaccines have become available, but effective treatments that reduce mortality are scarce, mainly in hospitalized or critically ill patients. Considering the pathophysiology of the disease and the need for a rapid response to the emergency, several treatments have been proposed, especially repurposing drugs with anti-inflammatory or antiviral action.^1,2^ Up to now, dexamethasone and tocilizumab has shown to decrease 28-day mortality among patients on mechanical ventilation or supplemental oxygen.^3,4^ Other drugs initially proposed (hydroxychloroquine, azithromycin, lopinavir/ritonavir, interferon beta, EMT) have no demonstrated benefit and, for others (remdesivir, tofacitinib, and baricitinib plus remdesivir), clinical impact on mortality is still not clear.^5–9^

Other drugs that have been proposed as potential treatment options for Covid-19 are statins, colchicine, and tenofovir/emtricitabine. Statins, mainly due to their anti-inflammatory, immunomodulatory and antithrombotic properties, have been considered to have a potential effect against Covid-19-associated complications.^10,11^ Colchicine has a rapid anti-inflammatory effect, by inhibiting NLRP3 inflammasome activation, neutrophil chemotaxis, and superoxide radical production, potentially preventing cytokine storm development.^12,13^ Finally, tenofovir and emtricitabine, nucleotide analogs widely used as antiretroviral therapy in HIV, potentially inhibit Severe Acute Respiratory Syndrome Coronavirus 2 (SARS-CoV-2) ribonucleic acid (RNA)-dependent RNA polymerase, which plays a fundamental role in transcription and viral replication.^14,15^

Although clinical information on the impact of the administration of each of these drugs to Covid-19 patients is already available from observational studies, results are not yet conclusive.^16–18^ In addition, there is little information from randomized studies on the combination of these therapeutic options in critically ill patients with Covid-19.^19,20^ Therefore, we proposed to evaluate the effectiveness of the combination of rosuvastatin plus colchicine, searching for an additional benefit from their combined use, the effects of emtricitabine/tenofovir disoproxil and, finally, the combined effect of the antiviral plus the anti-inflammatory drugs. These groups were compared to the standard of care in hospitalized Covid-19 patients, within the context of an open-label randomized clinical trial.

## METHODS

### Trial design and oversight

This is a pragmatic open parallel-group multi-centre randomised controlled trial. The protocol was previously published in Clinicaltrials.gov under the NCT04359095 identifier. The study protocol and informed consent template were approved by the Universidad National de Colombia Ethics Committee and by the Institutional Review Boards of the participating hospitals (protocol’s details are provided in the Supplementary Material). The study was funded by the Colombian Ministry of Science and Technology and coordinated by the Clinical Research Institute at the Universidad Nacional de Colombia; the generation of the random allocation sequence and the statistical analysis was carried out by the Department of Clinical Epidemiology and Biostatistics at the Pontificia Universidad Javeriana.

### Study population

Adults aged 18 years or more, with a positive real-time polymerase chain reaction (RT-PCR) or with high suspicion of SARS CoV-2 by clinical criteria and a diagnosis of mild, severe, or critical pneumonia, requiring hospital management in six high complexity referral hospitals located in Bogota.

Mild pneumonia was defined by chest X-ray images and 2 or more risk factors for Covid-19 complications, including age over 60 years, previous cardiovascular disease, diabetes mellitus, chronic obstructive pulmonary disease (COPD), or cancer. Moderate pneumonia was defined by chest X-ray images and hospitalization criteria of the simplified severity scale CRB-65>1 or oxygen saturation at ambient air of less than 90%. Severe pneumonia was defined with the same criteria as for moderate pneumonia, plus respiratory rate over 30 breaths per minute, or need for mechanical ventilation (invasive or non-invasive), or sepsis identified by a Sequential Organ Failure Assessment (SOFA) score of two or more points or quick SOFA with 2 out of 3 clinical variables, namely, Glasgow 13 or lower, systolic pressure of 100 mmHg or lower and respiratory rate of 22 breaths per minute or higher; or with a diagnosis of Septic Shock or Multiple organ failure or Adult Respiratory Distress Syndrome. Pregnant women, patients taking any of the study medications in the last 7 days or with known allergy to them, or with a history of myopathy or rhabdomyolysis, hepatic, or renal failure or lung fibrosis, advanced or metastatic cancer, and those with score more than 3 on the frailty scale, were excluded.

### Procedure

At each hospital, two general practitioners looked for candidates to include in the study. After verifying inclusion and exclusion criteria, the patients were invited to participate and to sign the informed consent. If a patient was unable to receive the information, a relative and a witness were asked to sign the informed consent on behalf of the eligible patient and to provide baseline information. Once the informed consent was filled out, data were recorded using a Web-based secure electronic database (Research Electronic Data Capture [REDCap], Ver. 6.16 Vanderbilt University). Information about baseline sociodemographic characteristics, previous comorbidities, functional condition at entry and level of respiratory support, were collected.

### Randomization

Patients were then randomly assigned to one of the four arms using a Web-based randomization system (UNcovApp ®) that allows maintaining concealment until the closest time to the start of treatment in one of the 4 arms. In each hospital, eligible and consenting patients were assigned in a 1:1 ratio to receive: a) usual standard of care or b) emtricitabine (200 mg) + tenofovir disoproxil (300 mg), once a day for 10 days PO, or c) colchicine, 0.5 mg twice a day, plus rosuvastatin, 40 mg once a day for 14 days PO, or d) emtricitabine/tenofovir disoproxil + colchicine + rosuvastatin at the same doses and duration, orally. The standard of care follows the recommendations of the Colombian Consensus for Covid-19 Treatment in Hospitalized Patients,^21^ consisting of the use of dexamethasone, antiparasitic treatment (ivermectin or albendazole), enoxaparin, acetaminophen, oxygen as needed, and supportive treatment for organ failure (i.e., use mechanical ventilation, or dialysis). The assigned treatment was prescribed by the treating physician in each institution. Patients and local researchers who evaluated the outcomes were aware of this information.

Patients were followed daily until discharge from the hospital, death, or at day 28. Information about adherence to treatments was entered in the UNcovApp® database. Information about clinical course, clinical status, requirement of respiratory or other organ function support, laboratory results and the primary and secondary outcomes ware recorded in the Web-based case report form. Patients who were discharged were followed by phone on post-assignment days 7 and 28. No information was gathered after day 28.

### Outcomes Primary outcomes

The primary effectiveness outcome was all-cause mortality within 28 days after randomization. Severe adverse events were those with a high probability (more than 50%) of being related to the assessed drugs or the standard treatment or organ function support devices (i.e., mechanical ventilator). Secondary outcomes were: 7-day mortality, proportion of intensive care unit (ICU) admission, proportion of mechanical respiratory support requirement, time to death, length of stay, and any adverse event related to the assessed treatments. Definitive discontinuation of one medication after 24 hours of treatment was considered as non-adherence.

### Analysis

We estimated that a sample size of 814 patients (204 per treatment arm) would provide the trial with 80% power to detect an absolute difference of 10% compared with the standard of care arm in the incidence of the primary outcome, assuming that 5% of the participants in the treatment groups and 15% of those in the standard of care group would have an event (*i*.*e*., death through day 28).^22,23^ The hypothesis of no difference was tested at a two-tailed alpha level of 5%. Additionally, we calculated a scenario with a sample size of 686 patients (172 per arm) using an alpha level of 10%, considering the absence of an effective treatment, beyond dexamethasone, in patients with moderate or severe pneumonia, and the pragmatic approach.^24^ On the other hand, the consequences for patients with Covid-19 of making a Type II error (concluding that some treatment is not effective when, in fact, it is) are far more costly than those of making a Type I error (concluding that any of study treatments is effective when, in fact, it is not). The last scenario was used as a stopping rule if we were unable to achieve the sample size in the first scenario. In both scenarios, sample sizes were adjusted based on Bonferroni multiple comparisons (3, each treatment versus standard treatment) and 10% withdrawals (See protocol in Supplementary Material).

Statisticians were not aware of the treatment received by patients. It was codified to maintain masking to them.

A descriptive analysis of the baseline conditions and adverse events by arm of treatment was carried out. Relative risk (RR) and risk difference (RD) with 95% confidence interval (95% CI) were estimated for 28-day mortality, 7-day mortality, ICU admission and required invasive mechanical ventilation outcomes using Log-binomial General Estimating Equation (GEE), assuming exchangeable correlation structure with each center as a cluster.

Unadjusted and adjusted by age, sex and severity of pneumonia models were performed for all outcomes. Standard error for RD was estimated by the delta method. Also, we estimated a crude RR and RD assuming independence between observations. A long-rank test was performed for time to death and length of stay. Kaplan–Meier survival curves were calculated to show cumulative mortality over the 28 days. Intention to treat analysis was done. On the other hand, as part of the pragmatic approach, the ranking of the treatment effect was estimated by an empirical bootstrap with 10,000 replicates and the surface under the cumulative ranking curve (SUCRA) was calculated for each arm.^25^

## RESULTS

### Patients

Between August 24, 2020, and March 20, 2021, a total of 1576 patients were seen in the six hospitals; 599 (38%) had some exclusion criteria, mainly chronic use of statins in 582.

Overall, 994 patients were invited to participate in the study, 328 (33%) refused to sign the informed consent, 649 were randomized and assigned to one of four arms of treatment. Of them, 3 subjects did not meet protocol selection criteria, 3 subjects were lost of follow-up and 10 patients withdrew; consequently, the primary outcome was unknown in 13 patients, so they were not included in the analysis. Finally, 633 patients were considered in the modified ITT analysis (Figure 1).^26^

**Figure 1.**
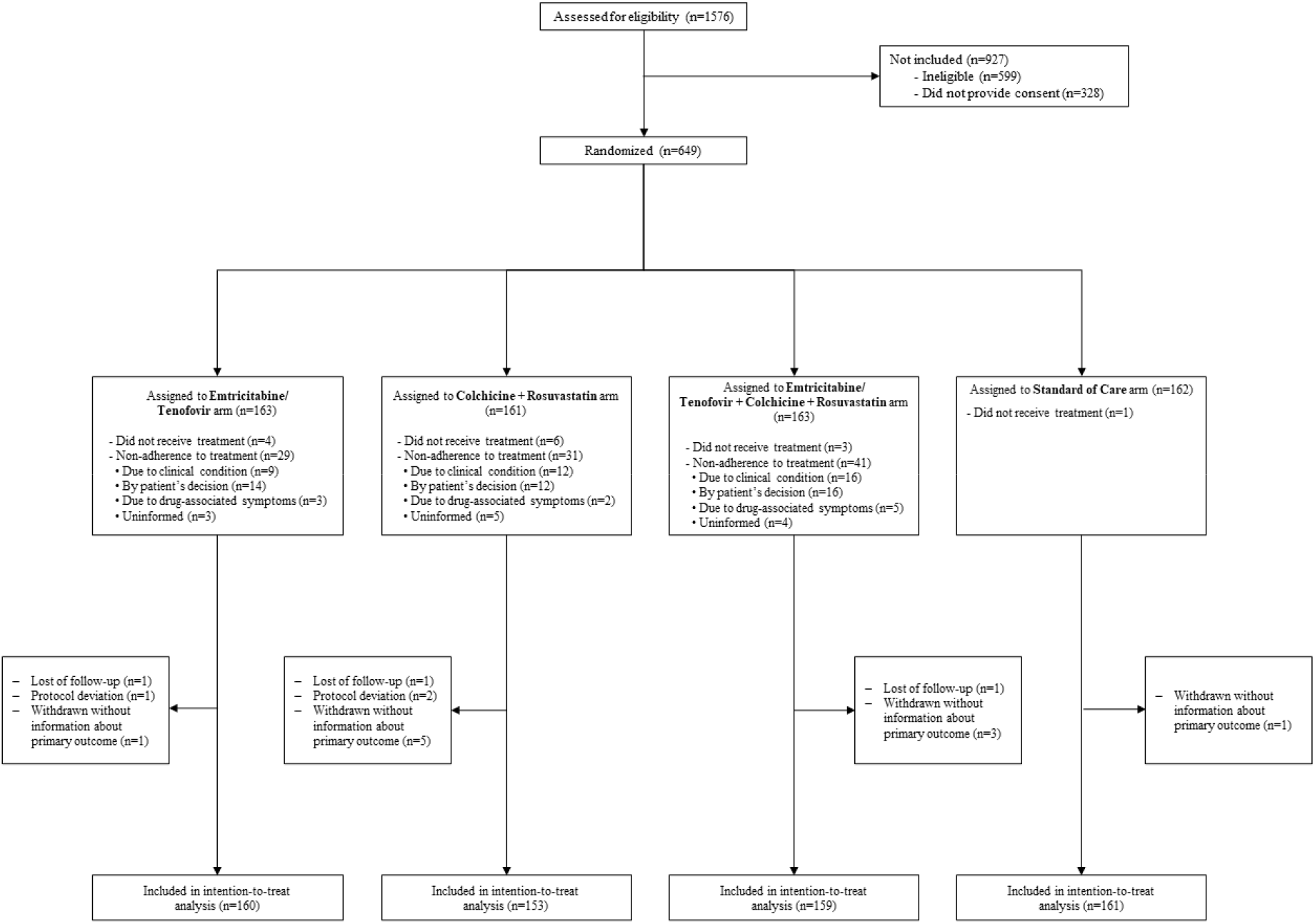
Enrollment, Randomization, and Inclusion in the intention-to-treat analysis. 1576 patients were seen in the six hospitals; 599 (38%) had some exclusion criteria, mainly chronic use of statins in 582. Overall, 994 patients were invited to participate in the study, 328 (33%) refused to sign the informed consent, 649 were randomized and assigned to one of four arms of treatment. Of them, 3 subjects did not meet protocol selection criteria, 3 subjects were lost of follow-up and 10 patients withdrew; consequently, the primary outcome was unknown in 13 patients, so they were not included in the analysis. Finally, 633 patients were considered in the modified ITT analysis.

The mean (± standard deviation) age of the participants was 55.4±12.8 years, and 32% were female. A history of diabetes was present in 12%, hypertension in 28%, COPD in 4%, and tobacco smoking in 16.2% of patients. At randomization, 7% were classified as mild pneumonia, 67% as moderate and 26% as severe, 8% were on invasive mechanical ventilation, 9% were receiving high flow nasal cannula, 66% oxygen only without invasive ventilation, and 17% were receiving neither (Table 1). Full adherence to the assigned treatment in the intervention groups varies between 18% and 25%; the distributions and causes are described in Figure 1. Patients did not receive other antiviral or anti-inflammatory treatments beyond those described above as standard treatment, and 98% of patients received dexamethasone across the 4 arms (Table S5).

**Table 1.** Characteristics of the Patients at Baseline, According to Treatment Assignment

| Characteristic * | Treatment assignment |  |  |  |
| --- | --- | --- | --- | --- |
|  | Emtricitabine/<br>Tenofovir | Colchicine<br>+<br>Rosuvastatin | Emtricitabine/<br>Tenofovir<br>+ Colchicine<br>+ Rosuvastatin | Standard of<br>Care |
|  | (N=160) | (N=153) | (N=159) | (N=161) |
| Female sex — no. (%) | 60 (37.5) | 50 (32.7) | 47 (29.6) | 48 (29.8) |
| Age |  |  |  |  |
| Mean age±SD — yr | 56.6±13.1 | 56.1±13.2 | 53.6±12.6 | 55.3±12.3 |
| Over 60 years — no. (%) | 66 (41.2) | 62 (40.5) | 47 (29.6) | 61 (37.9) |
| Urban Residency — no. (%) | 152 (96.8) | 149 (98.0) | 152 (96.8) | 156 (96.9) |
| Marital status — no. (%) |  |  |  |  |
| Single | 25 (16.2) | 21 (13.9) | 29 (18.5) | 24 (15.4) |
| Married | 91 (59.1) | 99 (65.6) | 94 (59.9) | 94 (60.3) |
| Other | 38 (24.7) | 31 (20.5) | 34 (21.7) | 38 (24.4) |
| Schooling — no. (%) |  |  |  |  |
| Primary or less | 13 (8.6) | 17 (11.9) | 13 (8.6) | 20 (13.2) |
| Secondary or Middle | 54 (35.5) | 52 (36.4) | 48 (31.6) | 47 (30.9) |
| Technical and technological | 22 (14.5) | 19 (13.3) | 18 (11.8) | 17 (11.2) |
| Professional and specialized | 63 (41.4) | 55 (38.5) | 73 (48.0) | 68 (44.7) |
| Social stratification (%) † |  |  |  |  |
| Medium-Low | 110 (71.0) | 110 (74.3) | 102 (68.0) | 102 (65.8) |
| Medium-High | 45 (29.0) | 38 (25.7) | 48 (32.0) | 53 (34.2) |
| Previous coexisting conditions — no. (%) |  |  |  |  |
| Smoking | 24 (15.3) | 21 (14.2) | 26 (16.9) | 33 (20.9) |
| Alcoholism | 7 (4.5) | 3 (2.0) | 4 (2.6) | 7 (4.5) |
| Cardiovascular disease | 9 (5.6) | 4 (2.6) | 2 (1.3) | 2 (1.2) |
| Diabetes mellitus 1-2 | 21 (13.1) | 24 (15.7) | 19 (11.9) | 12 (7.5) |
| Chronic respiratory disease | 9 (5.6) | 6 (3.9) | 5 (3.1) | 8 (5.0) |
| Arterial hypertension | 46 (28.7) | 51 (33.3) | 50 (31.4) | 29 (18.0) |
| Cancer | 9 (5.6) | 8 (5.2) | 6 (3.8) | 5 (3.1) |
| Obesity | 58 (38.2) | 48 (34.0) | 58 (39.2) | 45 (30.2) |
| Glomerular Filtration Rate — mean±SD | 88.3±19.4 | 91.7±17.4 | 91.4±19.6 | 92.8±16.6 |
| Length of stay in previous month — no. (%) | 0 (0.0) | 3 (2.0) | 3 (1.9) | 7 (4.4) |
| Charlson CCI — mean±SD | 0.3±0.7 | 0.4±0.9 | 0.3±0.7 | 0.2±0.6 |
| NEWS2 Classification — no. (%) |  |  |  |  |
| Low | 66 (41.2) | 59 (38.6) | 69 (43.7) | 74 (46.0) |
| Medium | 55 (34.4) | 54 (35.3) | 55 (34.8) | 51 (31.7) |
| High | 39 (24.4) | 40 (26.1) | 34 (21.5) | 36 (22.4) |
| Epidemiologic contact history — no. (%) |  |  |  |  |
| Known contact history | 58 (36.2) | 53 (34.6) | 63 (39.6) | 66 (41.0) |
| Community contact | 55 (94.8) | 50 (94.3) | 57 (90.5) | 60 (90.9) |
| Transportation contact | 0 (0.0) | 1 (1.9) | 0 (0.0) | 1 (1.5) |
| Healthcare staff | 3 (5.2) | 2 (3.8) | 6 (9.5) | 5 (7.6) |
| Service/Unit of current stay — no. (%) |  |  |  |  |
| General ward | 112 (70.0) | 109 (71.2) | 110 (69.2) | 114 (70.8) |
| ICU-Intensive | 44 (27.5) | 37 (24.2) | 44 (27.7) | 44 (27.3) |
| UCI-Step-down | 4 (2.5) | 7 (4.6) | 5 (3.1) | 3 (1.9) |
| Pneumonia diagnosis — no. (%) |  |  |  |  |
| Mild | 11 (6.9) | 12 (7.8) | 14 (8.8) | 8 (5.0) |
| Moderate | 109 (68.1) | 101 (66.0) | 102 (64.2) | 113 (70.2) |
| Severe | 40 (25.0) | 40 (26.1) | 43 (27.0) | 40 (24.8) |
| Median of days since onset of symptoms (Q1-Q3) | 9 (7-12) | 10 (6-12) | 10 (7-12) | 10 (7-12) |
| Median of days since admission (Q1-Q3) | 2 (1-3) | 2 (1-3) | 2 (1-3) | 2 (1-3) |
| Sepsis — no. (%) | 27 (65.9) | 24 (60.0) | 29 (67.4) | 28 (70.0) |
| Septic shock — no. (%) | 9 (22.5) | 8 (20.0) | 9 (20.9) | 10 (25.0) |
| ARDS — no. (%) | 39 (97.5) | 39 (97.5) | 43 (100.0) | 39 (97.5) |
| ARDS classification — no. (%) |  |  |  |  |
| Mild | 0 (0.0) | 1 (2.6) | 0 (0.0) | 1 (2.6) |
| Moderate | 19 (48.7) | 18 (46.2) | 14 (32.6) | 13 (33.3) |
| Severe | 20 (51.3) | 20 (51.3) | 29 (67.4) | 25 (64.1) |
| Oxygen delivery method — no. (%) |  |  |  |  |
| Non-invasive support | 100 (62.5) | 103 (67.3) | 107 (67.3) | 108 (67.1) |
| High-flow cannula | 16 (10.0) | 12 (7.8) | 13 (8.2) | 16 (9.9) |
| Mechanical ventilation | 14 (8.8) | 12 (7.8) | 12 (7.5) | 11 (6.8) |
| No oxygen | 30 (18.8) | 26 (17.0) | 27 (17.0) | 26 (16.1) |

\* Plus–minus values are means ± SD, ICU intensive care unit, ARDS Acute Respiratory Distress Syndrome, SD Standard Deviation, and Q1-Q3 25th percentile and 75th percentile.
† Social stratification is a government classification that groups areas of residential property where the study subjects live. Under this classification, Medium-Low corresponds to subject who benefit from utility subsidies and Medium-High are subjects who either do not receive subsidies and do not have to pay contributions or must pay a premium on the cost of utilities. DANE, *La estratificación socioeconómica en el régimen de los servicios públicos domiciliarios*, retrieved on June 16, 2021 at https://www.dane.gov.co/files/geoestadistica/Estratificacion_en_SPD.pdf

### Primary Outcome

Mortality at 28 days was not different between the intervention groups and the standard of care group (Table 2), the emtricitabine/tenofovir + colchicine + rosuvastatin arm reported 17 of 159 patients (10.7%), the colchicine + rosuvastatin arm reported 22 of 153 patients (14.4%), the emtricitabine/tenofovir arm reported 22 of 160 patients (13.8%), and the standard of care arm 28 of 161 patients (17.4%), with adjusted RD (aRD) against the standard of care arm of -0.07 (95% CI: -0.17 to 0.04), aRD -0.03 (95% CI: -0.11 to 0.05) and aRD -0.05 (95% CI: -0.15 to 0.05), respectively. Sensitivity analyses for the primary outcome, involving analysis with unadjusted estimates and analysis including patients with loss of follow-up, showed results similar to those of the main analysis (Table S1, S2 and S3). In an exploratory analysis by severity of pneumonia, the emtricitabine/tenofovir + colchicine + rosuvastatin arm decreased the 28-day mortality compared with the standard of care arm in patients with mild-moderate pneumonia (Table S4). The ranking of the aRD from SUCRA analysis also showed that the emtricitabine/tenofovir + colchicine + rosuvastatin arm had the highest probability to be the best option of the four treatments (SUCRA 76.2%) (Figure 2).

**Table 2.** Primary and Secondary Outcomes with Adjusted Estimates

| Outcomes * | No. Events /Total of patients (%) | Adjusted Relative Risk § (95% CI) | Adjusted Risk Difference § (95% CI) |
| --- | --- | --- | --- |
| **28-day mortality (N=633)** |  |  |  |
| *Arms* |  |  |  |
| Colchicine + Rosuvastatin | 22/153 (14.4) | 0.81 (0.48 to 1.35) | -0.03 (-0.11 to 0.05) |
| EMT/TNF + Colchicine + Rosuvastatin | 17/159 (10.7) | 0.62 (0.33 to 1.18) | -0.07 (-0.17 to 0.04) |
| EMT/TNF | 22/160 (13.8) | 0.70 (0.38 to 1.32) | -0.05 (-0.15 to 0.05) |
| Standard of Care | 28/161 (17.4) |  |  |
| **7-day mortality (N=633)** |  |  |  |
| *Arms* |  |  |  |
| Colchicine + Rosuvastatin | 9/153 (5.9) | 1.23 (0.26 to 5.77) | 0.01 (-0.06 to 0.08) |
| EMT/TNF + Colchicine + Rosuvastatin | 6/159 (3.8) | 0.81 (0.29 to 2.25) | -0.01 (-0.06 to 0.04) |
| EMT/TNF | 7/160 (4.4) | 0.91 (0.28 to 2.98) | 0.00 (-0.06 to 0.05) |
| Standard of Care | 7/161 (4.3) |  |  |
| **Transfer to the ICU (N=452) †** |  |  |  |
| *Arms* |  |  |  |
| Colchicine + Rosuvastatin | 19/113 (16.8) | 0.99 (0.56 to 1.74) | 0.00 (-0.09 to 0.09) |
| EMT/TNF + Colchicine + Rosuvastatin | 14/112 (12.5) | 0.87 (0.69 to 1.10) | -0.02 (-0.06 to 0.01) |
| EMT/TNF | 19/113 (16.8) | 0.92 (0.41 to 2.09) | -0.01 (-0.14 to 0.12) |
| Standard of Care | 18/114 (15.8) |  |  |
| **Ventilation requirement (N=556) ‡** |  |  |  |
| *Arms* |  |  |  |
| Colchicine + Rosuvastatin | 27/136 (19.9) | 0.85 (0.68 to 1.06) | -0.03 (-0.07 to 0.01) |
| EMT/TNF + Colchicine + Rosuvastatin | 20/141 (14.2) | 0.70 (0.52 to 0.95) | -0.06 (-0.11 to -0.01) |
| EMT/TNF | 22/139 (15.8) | 0.65 (0.37 to 1.12) | -0.07 (-0.16 to 0.01) |
| Standard of Care | 27/140 (19.3) |  |  |

\* ICU intensive care unit, and EMT/TNF Emtricitabine/Tenofovir.
† Patients who were in the ICU at the time of randomization are excluded.
‡ Patients who required ventilation before randomization are excluded.
§ Adjusted Relative Risk and Adjusted Risk Difference have been adjusted for age, sex, and severity of pneumonia using Log-binomial General Estimating Equation models, assuming exchangeable correlation structure with each center as a cluster.

**Figure 2.**
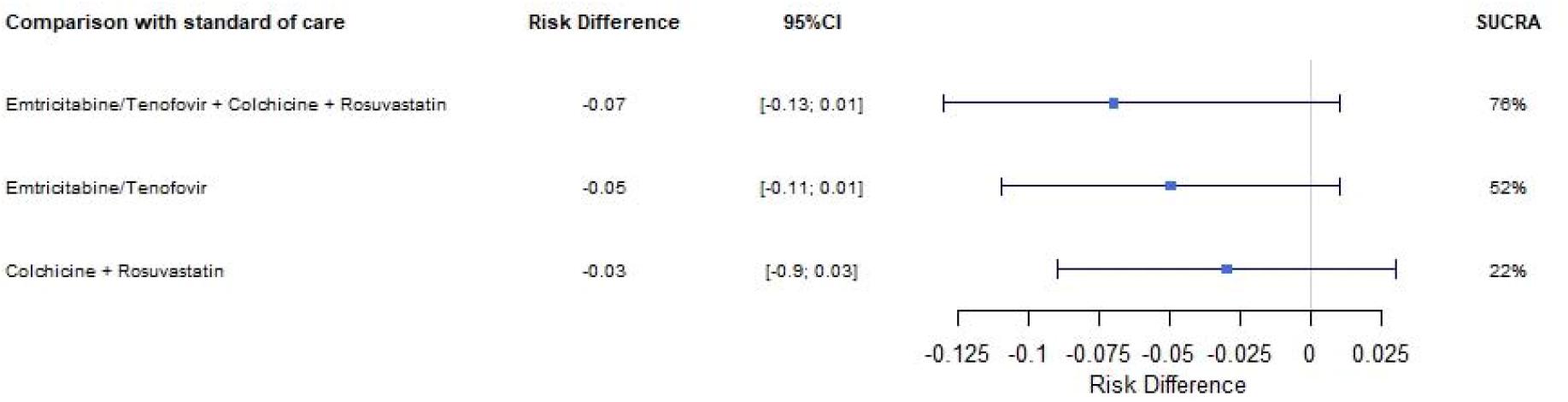
Surface Under the Cumulative Ranking Curve (SUCRA) analysis. Estimated adjusted risk differences in mortality between treatments and standard of care and 95% CI by 10.000 bootstrap replications. Sex, age, and severity of pneumonia were used on the adjustment. SUCRA represents the overall ranking; the higher value represents a higher likelihood that the treatment will be one of the treatments with the highest differences with the standard of care after adjustment.

Regarding safety, 3 patients with SAEs were identified, 1 patient in the emtricitabine/tenofovir + colchicine + rosuvastatin arm (severe diarrhea), 1 in the colchicine + rosuvastatin arm (severe diarrhea), and 1 patient in the emtricitabine/tenofovir arm (drug-associated exanthema).

### Other pre-specified secondary outcomes

The need for invasive mechanical ventilation after treatment assignment was lower in the emtricitabine/tenofovir + colchicine + rosuvastatin arm that reported 20 of 141 patients (14.2%) compared to the standard arm that reported 27 of 140 patients (19.3%) with an aRD of -0.06 (95% CI: -0.11 to -0.01). No differences were found between the colchicine + rosuvastatin arm or the emtricitabine/tenofovir arm, and the standard treatment (Table 2).

There were no differences in mortality at 7 days and transfer to the ICU between the treatment arms and the standard of care group (Table 2), neither in terms of median time to death between the colchicine + rosuvastatin arm (9.0 days, 25th percentile and 75th percentile [Q1-Q3]: 4.0-12.0), emtricitabine/tenofovir arm (12.0 days, Q1-Q3: 7.0-15.0), emtricitabine/tenofovir + colchicine + rosuvastatin arm (12.0 days, Q1-Q3: 7.0-18.0), and standard of care arm (13.5 days, Q1-Q3: 7.5-18.5) (Log-rank test for equality of survivor functions, p-value:0.41), or the medians (Q1-Q3) for length of stay between colchicine + rosuvastatin arm (4.0 days (2.0-9.0)), emtricitabine/tenofovir arm (5 days (3-10)), emtricitabine/tenofovir + colchicine + rosuvastatin arm (5.0 days (2.5-8.0)) and standard of care (4.0 days (2.0-9.0)). The more frequent non-serious adverse events are shown in Table 3.

**Table 3.** Frequency of non-serious adverse events

| **Description of non-serious adverse events \*** | **Total** | **Emtricitabine/**<br>**Tenofovir** | **Colchicine**<br>**+**<br>**Rosuvastatin** | **Emtricitabine/**<br>**Tenofovir**<br>**+ Colchicine**<br>**+ Rosuvastatin** | **Standard**<br>**of Care** |
| --- | --- | --- | --- | --- | --- |
| Gastrointestinal (nausea, diarrhea, epigastralgia) | 88 | 22 | 24 | 38 | 4 |
| Hepatic (elevation of transaminases, alkaline phosphatase, and bilirubin) | 67 | 15 | 17 | 24 | 11 |
| Non-specific (asthenia, cramps, diaphoresis) | 23 | 6 | 6 | 9 | 2 |
| Neurologic (headache, delirium, seizure episode) | 22 | 5 | 2 | 10 | 5 |
| Cardiovascular (hypertension, bradycardia, atrial fibrillation) | 21 | 6 | 4 | 6 | 5 |
| Renal (kidney injury, hematuria, and increased creatine phosphokinase) | 20 | 7 | 7 | 1 | 5 |
| Hematologic (Anemia, thrombocytopenia, thrombocytosis) | 16 | 2 | 5 | 5 | 4 |
| Allergic (Exanthema, rash, and allergic reaction) | 14 | 5 | 6 | 3 | 0 |
| Metabolic (diabetes, hyperglycemia and cholelithiasis) | 12 | 6 | 2 | 3 | 1 |
| Osteomuscular (contracture, weakness, myalgia) | 6 | 2 | 1 | 3 | 0 |
| Infectious (bacteremia, herpes zoster and catheter site infection) | 5 | 1 | 2 | 0 | 2 |
| Electrolytes (Hyperkalemia and hypokalemia) | 4 | 1 | 2 | 0 | 1 |
| Psychiatric (Anxiety, panic, and psychosis) | 3 | 1 | 0 | 1 | 1 |
| Respiratory (Dyspnea and pneumothorax) | 3 | 1 | 2 | 0 | 0 |
| Otolaryngological (Otorrhagia) | 2 | 1 | 1 | 0 | 0 |
| Dental (Tooth loss) | 1 | 0 | 1 | 0 | 0 |

\* We do not present information with relative frequencies (i.e., percentages) because some events occurred simultaneously in the same patients; therefore, it is difficult to have a denominator.

## DISCUSSION

This pragmatic randomized trial shows that, in hospitalized patients with Covid-19 and mild, moderate, or severe pneumonia, the use of emtricitabine/tenofovir disoproxil together with colchicine and rosuvastatin reduces the need for invasive mechanical ventilation by 6.0% in comparison with the standard of care. Regarding the primary outcome, a clinically significant trend towards lower 28-day mortality was found, with an adjusted risk difference of -6.7%. However, this result is imprecise because the required sample size was not achieved. The results were consistent with the crude analysis and the unadjusted analysis using GEE, when patients with mild-moderate or severe pneumonia were considered separately, and in the scenario including patients with incomplete data. In the arms of our study, mortality falls within the range described for Western countries.^5,27^

Our findings support the potential benefits of combined antiviral and anti-inflammatory therapies based on the current knowledge of the pathophysiology of SARS-CoV-2. This combination seeks to decrease the replication capacity and cytopathic damage of Covid-19, as well to control the overactivation of the innate immune system, the hyperinflammatory state and the consequent endothelial dysfunction, cell injury and multi-organ damage.^28,29^ The proposed combination could produce a more significant synergistic effect than the use of each medication alone.

In the case of colchicine, the current evidence is conflicting. The COLCORONA study, a masked RTC that included 4488 Covid-19 outpatients, showed a reduction in the incidence of the combined primary outcome of death or hospitalization compared to placebo (Odds Ratio [OR] 0.75, 95%CI: 0.57 to 0.99).^30^ Benefit was also reported by the GRECO study authors in 105 hospitalized patients with Covid-19 in which patients treated with colchicine showed a reduction of the time to clinical deterioration (OR 0.11, 95% CI 0.01 to 0.96).^19^ However, the RECOVERY trial with 2363 hospitalized Covid-19 patients showed that colchicine was not associated with mortality reduction (RR 1.01; 95% CI 0.93 to 1.10), but they do not report data of invasive ventilation requirement alone.^31^ Regarding the use of statins in the treatment of Covid-19, although 22 RCTs are now registered in clinicaltrials.gov and reported as completed, we could not find any published study to date. So far, evidence about its benefits in patients with Covid-19 comes from observational studies.^32,33^ The Vahedian-Azimi meta-analysis that included 24 studies with 32,715 patients, showed significant reductions in ICU admission (OR 0.78, 95% CI: 0.58 to 1.06) and death (OR 0.70, 95% CI: 0.55 to 0.88). Interestingly a subgroup analysis suggested that death was reduced further by in-hospital administration of statins compared to pre-hospital use.^33^ Regarding tenofovir/emtricitabine, 8 studies were found in clinicaltrials.gov, but none have been completed.

Our study has some limitations. The number of patients included was lower than expected, affecting the precision of our results. Low recruitment was due partly to a high proportion of patients who refused to provide their consent to participate (33%), a cultural issue that should be analyzed. Also, the lack of additional funding prevented us from continuing the study and, it had to be stopped based on the second sample size scenario. Another problem was detection bias, as it was an open study. The effect is found in the proportion of non-adherence to treatment, ranging between 18% and 25% in the different study medication groups (Figure1). Considering various scenarios, the direction of this bias could favor the null hypothesis; consequently, the effect in terms of mortality reduction with the combined treatment could be accurate. In terms of generalizability, a significant number of patients on chronic treatment with statins were considered not eligible (37%) based on the exclusion criteria. Therefore, our findings are only relevant for patients in whom the use of statins is novel, which affects the pragmatic approach of the study. Moreover, the study was conducted under the usual conditions in hospitals (with high workloads and standard medication administration systems at each hospital), using an ITT approach analysis, and looking for the more useful treatment against the standard of care to help the clinical decision analysis.

## CONCLUSIONS

The combined use of emtricitabine/tenofovir disoproxil + rosuvastatin + colchicine for 14 days emerges as an option to consider in the treatment of patients with moderate and severe Covid-19, based on its effects of reducing the use of invasive mechanical ventilation and a clinically significant reduction of 28-day mortality both in patients with moderate and severe pneumonia. Further RCTs that assess this combination are needed to validate our results and reduce imprecision in estimating its effect on mortality according to the severity of pneumonia or the level of respiratory dysfunction, and in patients with chronic use of statins.

## Data Availability

The data will be shared upon request to the corresponding author: Hernando Gaitan-Duarte

## Funding

This project was supported by the Grant 374-2020 given from the Ministerio de Ciencia y Tecnología de Colombia (Minciencias) to the Universidad Nacional de Colombia. Additionally, it was supported by Universidad Nacional de Colombia, Pontificia Universidad Javeriana, Clínica Colsanitas SA, Hospital Universitario San Ignacio, Fundación Cardio Infantil and Hospital Universitario Nacional de Colombia.

## Supplementary Appendix

## Appendix 1.

Acknowledgment. List of Hospitals and Investigators.

<u>Clínica Santa María del Lago:</u> Andrés Mauricio Gómez Saza, Diego Armando Bornachera Pinto, María de los Ángeles Cuellar Losada, Nicolás Campos Gómez, Diana Carolina Alfonso Vergel, Albert Valencia.

<u>Fundación Cardioinfantil-Instituto de Cardiología:</u> Eliana Yineth Vaquiro Herrera, Fabio Varón, Luis David Sáenz Prieto, Juan Sebastián Sánchez Solarte.

<u>Clínica Reina Sofía:</u> Viviana Lorena Martínez Pinzón, Dania Jimena Zuluaga Sierra, Jennifer Tatiana Aristizabal Robayo, Ángela Consuelo Díaz Sierra, Giovanna Katherine Lasso Araujo, Juliana María Rueda Garzón.

<u>Clínica Universitaria Colombia:</u> Jheymi Catalina Ramos Bohórquez, Daniela Aldana García, Andreina Martínez Amado, Kelly Rocío Chacón Acevedo, Diego Alejandro Pinto Pinzón, Carlos Eduardo Pinzón Flórez.

<u>Hospital Universitario Nacional:</u> Carmelo Espinosa, July Torres, Jairo Pérez Cely, Daniel Yamil Ariza Henríquez, Ana María Carreño G, Luis Armando Dulcey Cala.

<u>Hospital Universitario San Ignacio/ Pontificia Universidad Javeriana:</u> Margarita Manrique, María Julieta Pachón Espinosa, Oscar Muñoz, Daniela Alejandra Varela Herrera, Andrés Felipe Cárdenas Cruz.

<u>Universidad Nacional de Colombia:</u> Erika Tatiana Ruiz, Alejandro Troncoso, Mario Orlando Parra Pineda, Olga Janeth Gómez, Claudia Vaca González, Daniela Quintero, Gabriel Esteban González Espitia, Luis Miguel García Mora, Ángel Arturo Gómez Mora, Ruth Maldonado Silva.

We especially thank the hundreds of patients and their families who participated in this trial.

---

**Table S1.** Primary and Secondary Outcomes with Crude Estimates

| **Outcomes \*** | **No. Events /Total of patients (%)** | **Relative Risk § (95% CI)** | **Risk Difference § (95% CI)** |
| --- | --- | --- | --- |
| **28-day mortality (N=633)** |  |  |  |
| *Arms* |  |  |  |
| Colchicine + Rosuvastatin | 22/153 (14.4) | 0.83 (0.5 to 1.38) | -0.03 (-0.11 to 0.05) |
| EMT/TNF + Colchicine + Rosuvastatin | 17/159 (10.7) | 0.61 (0.35 to 1.08) | -0.07 (-0.14 to 0.01) |
| EMT/TNF | 22/160 (13.8) | 0.79 (0.47 to 1.32) | -0.04 (-0.12 to 0.04) |
| Standard of Care | 28/161 (17.4) |  |  |
| **7-day mortality (N=633)** |  |  |  |
| *Arms* |  |  |  |
| Colchicine + Rosuvastatin | 9/153 (5.9) | 1.35 (0.52 to 3.54) | 0.02 (-0.03 to 0.06) |
| EMT/TNF + Colchicine + Rosuvastatin | 6/159 (3.8) | 0.87 (0.3 to 2.53) | -0.01 (-0.05 to 0.04) |
| EMT/TNF | 7/160 (4.4) | 1.01 (0.36 to 2.8) | 0.00 (-0.04 to 0.04) |
| Standard of Care | 7/161 (4.3) |  |  |
| **Transfer to the ICU (N=452) †** |  |  |  |
| *Arms* |  |  |  |
| Colchicine + Rosuvastatin | 19/113 (16.8) | 1.06 (0.59 to 1.92) | 0.01 (-0.09 to 0.11) |
| EMT/TNF + Colchicine + Rosuvastatin | 14/112 (12.5) | 0.79 (0.41 to 1.51) | -0.03 (-0.12 to 0.06) |
| EMT/TNF | 19/113 (16.8) | 1.06 (0.59 to 1.92) | 0.01 (-0.09 to 0.11) |
| Standard of Care | 18/114 (15.8) |  |  |
| **Ventilation requirement (N=556) ‡** |  |  |  |
| *Arms* |  |  |  |
| Colchicine + Rosuvastatin | 27/136 (19.9) | 1.03 (0.64 to 1.66) | 0.01 (-0.09 to 0.1) |
| EMT/TNF + Colchicine + Rosuvastatin | 20/141 (14.2) | 0.74 (0.43 to 1.25) | -0.05 (-0.14 to 0.04) |
| EMT/TNF | 22/139 (15.8) | 0.82 (0.49 to 1.37) | -0.03 (-0.12 to 0.05) |
| Standard of Care | 27/140 (19.3) |  |  |

\* ICU intensive care unit, and EMT/TNF Emtricitabine/Tenofovir.
† Patients who were in the ICU at the time of randomization are excluded.
‡ Patients who required ventilation before randomization are excluded.
§ Crude Relative Risk and Risk Difference were estimated assuming independence between observations.

**Table S2.**
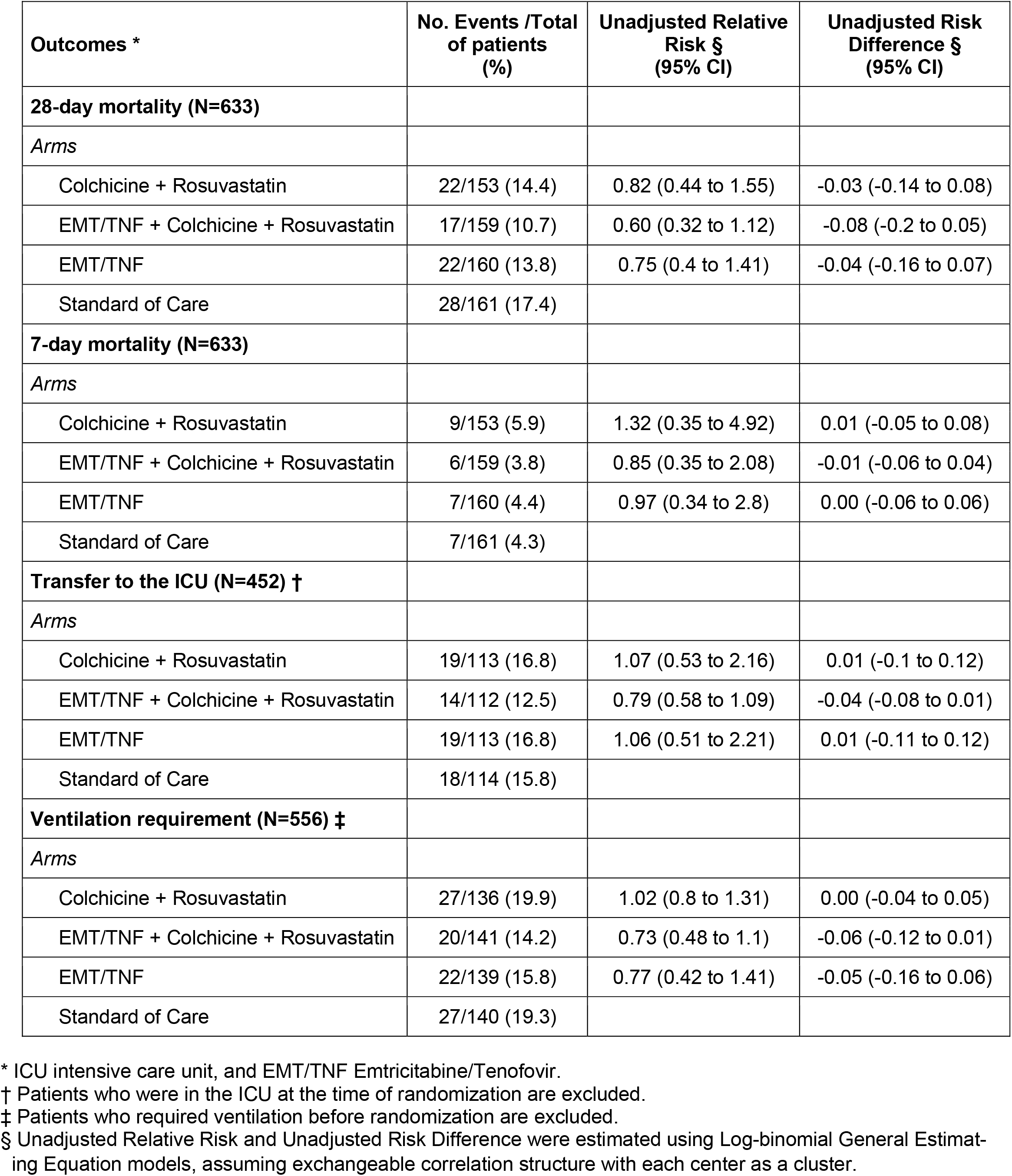
Primary and Secondary Outcomes with Unadjusted Estimates (GEE models)

| **Outcomes \*** | **No. Events /Total of patients (%)** | **Unadjusted Relative Risk § (95% CI)** | **Unadjusted Risk Difference § (95% CI)** |
| --- | --- | --- | --- |
| **28-day mortality (N=633)** |  |  |  |
| *Arms* |  |  |  |
| Colchicine + Rosuvastatin | 22/153 (14.4) | 0.82 (0.44 to 1.55) | -0.03 (-0.14 to 0.08) |
| EMT/TNF + Colchicine + Rosuvastatin | 17/159 (10.7) | 0.60 (0.32 to 1.12) | -0.08 (-0.2 to 0.05) |
| EMT/TNF | 22/160 (13.8) | 0.75 (0.4 to 1.41) | -0.04 (-0.16 to 0.07) |
| Standard of Care | 28/161 (17.4) |  |  |
| **7-day mortality (N=633)** |  |  |  |
| *Arms* |  |  |  |
| Colchicine + Rosuvastatin | 9/153 (5.9) | 1.32 (0.35 to 4.92) | 0.01 (-0.05 to 0.08) |
| EMT/TNF + Colchicine + Rosuvastatin | 6/159 (3.8) | 0.85 (0.35 to 2.08) | -0.01 (-0.06 to 0.04) |
| EMT/TNF | 7/160 (4.4) | 0.97 (0.34 to 2.8) | 0.00 (-0.06 to 0.06) |
| Standard of Care | 7/161 (4.3) |  |  |
| **Transfer to the ICU (N=452) †** |  |  |  |
| *Arms* |  |  |  |
| Colchicine + Rosuvastatin | 19/113 (16.8) | 1.07 (0.53 to 2.16) | 0.01 (-0.1 to 0.12) |
| EMT/TNF + Colchicine + Rosuvastatin | 14/112 (12.5) | 0.79 (0.58 to 1.09) | -0.04 (-0.08 to 0.01) |
| EMT/TNF | 19/113 (16.8) | 1.06 (0.51 to 2.21) | 0.01 (-0.11 to 0.12) |
| Standard of Care | 18/114 (15.8) |  |  |
| **Ventilation requirement (N=556) ‡** |  |  |  |
| *Arms* |  |  |  |
| Colchicine + Rosuvastatin | 27/136 (19.9) | 1.02 (0.8 to 1.31) | 0.00 (-0.04 to 0.05) |
| EMT/TNF + Colchicine + Rosuvastatin | 20/141 (14.2) | 0.73 (0.48 to 1.1) | -0.06 (-0.12 to 0.01) |
| EMT/TNF | 22/139 (15.8) | 0.77 (0.42 to 1.41) | -0.05 (-0.16 to 0.06) |
| Standard of Care | 27/140 (19.3) |  |  |

\* ICU intensive care unit, and EMT/TNF Emtricitabine/Tenofovir.
† Patients who were in the ICU at the time of randomization are excluded.
‡ Patients who required ventilation before randomization are excluded.
§ Unadjusted Relative Risk and Unadjusted Risk Difference were estimated using Log-binomial General Estimating Equation models, assuming exchangeable correlation structure with each center as a cluster.

**Table S3.** Sensitivity analysis of the main outcome (28-day mortality) including loss of follow-up

| Treatment arms * | No. Events /Total of patients (%) | Relative Risk (95% CI) | Risk Difference (95% CI) |
| --- | --- | --- | --- |
| *Crude Estimates* † |  |  |  |
| Colchicine + Rosuvastatin | 28/159 (17.6) | 0.98 (0.61 to 1.58) | 0.00 (-0.09 to 0.08) |
| EMT/TNF + Colchicine + Rosuvastatin | 21/163 (12.9) | 0.72 (0.43 to 1.21) | -0.05 (-0.13 to 0.03) |
| EMT/TNF | 24/162 (14.8) | 0.83 (0.50 to 1.36) | -0.03 (-0.11 to 0.05) |
| Standard of Care | 29/162 (17.9) |  |  |
| *Unadjusted Estimates (GEE models)* ‡ |  |  |  |
| Colchicine + Rosuvastatin | 28/159 (17.6) | 0.99 (0.44 to 2.22) | 0.00 (-0.14 to 0.14) |
| EMT/TNF + Colchicine + Rosuvastatin | 21/163 (12.9) | 0.71 (0.34 to 1.49) | -0.06 (-0.21 to 0.09) |
| EMT/TNF | 24/162 (14.8) | 0.80 (0.40 to 1.61) | -0.04 (-0.17 to 0.09) |
| Standard of Care | 29/162 (17.9) |  |  |
| *Adjusted Estimates (GEE models)* § |  |  |  |
| Colchicine + Rosuvastatin | 28/159 (17.6) | 0.90 (0.47 to 1.73) | -0.02 (-0.12 to 0.09) |
| EMT/TNF + Colchicine + Rosuvastatin | 21/163 (12.9) | 0.70 (0.33 to 1.48) | -0.05 (-0.18 to 0.07) |
| EMT/TNF | 24/162 (14.8) | 0.73 (0.36 to 1.47) | -0.05 (-0.16 to 0.07) |
| Standard of Care | 29/162 (17.9) |  |  |

\* 28-day mortality effects of the treatment arms compared with Standard of Care arm. The analysis of sensitivity included 646 patients (633 patients of main ITT analysis + 13 patients with loss of follow-up). We assumed that all 13 patients died during follow-up. EMT/TNF: Emtricitabine/Tenofovir.
† Crude Relative Risk and Risk Difference were estimated assuming independence between observations.
‡ Unadjusted Relative Risk and Unadjusted Risk Difference were estimated using Log-binomial General Estimating Equation models, assuming exchangeable correlation structure with each center as a cluster.
§ Adjusted Relative Risk and Adjusted Risk Difference have been adjusted for age, sex, and severity of pneumonia using Log-binomial General Estimating Equation models, assuming exchangeable correlation structure with each center as a cluster.

**Table S4.**
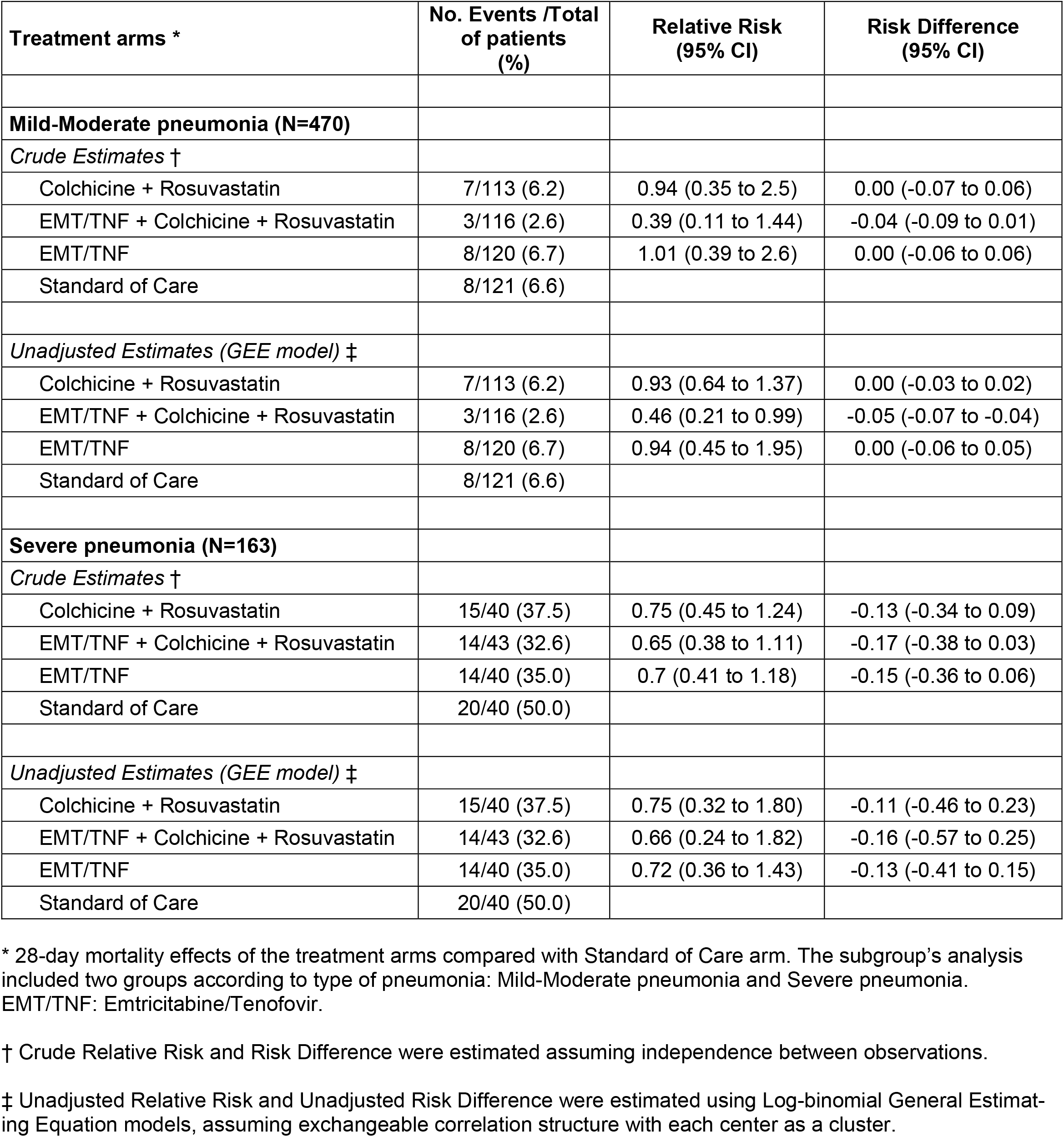
Subgroup’s analysis of the main outcome (28-day mortality) by severity of pneumonia

**Table S5.** Use of dexamethasone by treatment arm

| Treatment arms | Total of patients<br>by arm | Use of dexamethasone |  |
| --- | --- | --- | --- |
|  |  | Number | Proportion (%) |
| Colchicine + Rosuvastatin | 153 | 151 | 98.7 |
| EMT/TNF + Colchicine + Rosuvastatin | 159 | 154 | 96.9 |
| EMT/TNF | 160 | 157 | 98.1 |
| Standard of Care | 161 | 160 | 99.4 |

## Notes

### Competing Interest Statement

The authors have declared no competing interest.

### Clinical Trial

ClinicalTrials.gov number: NCT04359095

### Funding Statement

The study was funded by the Colombian Ministry of Science and Technology, Universidad Nacional de Colombia, and Pontificia Universidad Javeriana.

### Author Declarations

The study protocol and informed consent template were approved by the Universidad Nacional de Colombia Ethics Committee and by the Institutional Review Boards of the participating hospitals

